# Impact of regional heterogeneity on the severity of COVID-19

**DOI:** 10.1101/2021.06.30.21259758

**Authors:** Shinya Tsuzuki, Yusuke Asai, Nobuaki Matsunaga, Haruhiko Ishioka, Takayuki Akiyama, Norio Ohmagari

**Affiliations:** AMR Clinical Reference Center, National Center for Global Health and Medicine, Tokyo, Japan; Faculty of Medicine and Health Sciences, University of Antwerp, Antwerp, Belgium; Disease Control and Prevention Center, National Center for Global Health and Medicine, Tokyo, Japan

**Author notes:** Corresponding author: Shinya Tsuzuki, MD, MSc, National Center for Global Health and Medicine, 1-21-1 Toyama, Shinjuku-ku, Tokyo 162-8655, Japan.

**Keywords:** COVID-19, severity, regional heterogeneity, hierarchical Bayesian model

## Abstract

**Background:** We aimed to assess the impact of regional heterogeneity on the severity of COVID-19 in Japan.

**Methods:** We included 27,865 cases registered between January 2020 and February 2021 in the COVID-19 Registry of Japan to examine the relationship between the National Early Warning Score (NEWS) of COVID-19 patients on the day of admission and the prefecture where the patients live. A hierarchical Bayesian model was used to examine the random effect of each prefecture in addition to the patients’ backgrounds. In addition, we compared the results of two models; one model included the number of beds secured for COVID-19 patients in each prefecture as one of the fixed effects, and the other model did not.

**Results:** The results indicated that the prefecture had a substantial impact on the severity of COVID-19 on admission. Even when considering the effect of the number of beds separately, the heterogeneity caused by the random effect of each prefecture affected the severity of the case on admission.

**Conclusions:** Our analysis revealed a possible association between regional heterogeneity and increased/decreased risk of severe COVID-19 infection on admission. This heterogeneity was derived not only from the number of beds secured in each prefecture but also from other factors.

## 1. Introduction

Coronavirus disease 2019 (COVID-19), which is caused by the SARS-CoV-2 virus, has become a global health threat, imposing a substantial disease burden on our society.^1^ Although it has fluctuated, the epidemic has continued.^2^

COVID-19 is more infectious than other respiratory tract infections,^3^ such as influenza, and is sometimes fatal to the elderly and those with underlying medical conditions. Considering these characteristics, the capacity of healthcare facilities for COVID-19 patients is an important factor in the management of newly infected COVID-19 patients.

When we consider countermeasures against COVID-19, the proportion of patients with severe disease is extremely important.^4–6^ Mild and/or asymptomatic cases can be easily managed as they need no specific treatment. Conversely, severe cases often require intensive, multidisciplinary care. Furthermore, the duration of the disease in severe cases is generally longer than that of other viral pneumonias^7^ and therefore places a greater burden on healthcare capacity. It is important, therefore, to precisely recognize the factors associated with the severity of COVID-19. For example, new variants may contribute to the severity of COVID-19 cases.^8^ However, such variants are not the only determinants of severity; age, sex, past medical history, and many other factors have been attributed to the severity of COVID-19.^5,9,10^ In addition, it is conceivable that there are other factors influencing the severity of COVID-19 aside from such biological aspects.

In Japan, the number and proportion of severe COVID-19 cases vary by prefecture. This phenomenon seems difficult to explain; nevertheless, there are differences in the management of COVID-19 cases among prefectures, and these differences may contribute to the capacity of healthcare facilities in each prefecture. Understanding the extent of such regional differences will help us tailor the countermeasures against COVID-19 more appropriately in each prefecture. The main objective of our study was to examine this regional heterogeneity quantitatively.

## 2. Methods

We included 27,865 cases registered between January 2020 and February 2021 in the COVID-19 Registry of Japan (COVIREGI-JP)^11^. We used a Bayesian hierarchical model to construct a prefecture-specific random intercept to evaluate the effect of regional heterogeneity.

The capacity of medical facilities can be evaluated by the number of hospital beds; however, the differences in health policies against COVID-19 among local municipalities should also be taken into consideration as the planning of hospital bed distribution and the priority of hospitalization indication are influenced by the policy of each local municipality. Considering this, we constructed two models: one which included the number of beds secured for COVID-19 patients in each prefecture as a fixed effect (model B) and one which did not (model A).

The independent variable was the National Early Warning Score (NEWS)^12^ of COVID-19 patients on the day of admission, and the explanatory variables for the fixed effect were sex, age of 65 years and older, and any risk factor of severe infection (cardiovascular disease, cerebrovascular disease, chronic respiratory disease, liver disease, diabetes, hypertension, obesity, dialysis or sever renal failure, and malignancy) for model A and the number of beds secured for COVID-19 patients in each prefecture for model B.

The independent variable, the NEWS score at the time of admission, was assumed to follow a Poisson distribution. We set four separate sampling chains, each consisting of 12,000 random samples, including 1,000 burnin. The sampling convergence was evaluated using the Gelman–Rubin statistics (R-hat below 1.1) and by visually inspecting a trace plot. All analyses were conducted using the R software version 4.0.5.^13^

## 3. Results

Table 1 presents the demographic characteristics of the hospitalized COVID-19 cases registered in COVIREGI-JP. Tables 2 and 3 present the posterior distribution of the model parameters.

**Table 1.** Characteristics of registered patients.

| Variable | Number |
| --- | --- |
| Sex |  |
| Male | 16152 (58%) |
| Female | 11701 (42%) |
| Other | 9 (0%) |
| Days from symptom onset to hospitalization | 5 [2, 7] |
| Age | 57 [41, 73] |
| 0-64 | 16927 (60.7%) |
| 65- | 10938 (39.3%) |
| Ethnicity |  |
| Japanese | 26407 (95.8%) |
| Other ethnicity | 1156 (4.2%) |
| Smoking history |  |
| Former or current smoker | 10246 (36.9%) |
| Never | 12771 (46.0%) |
| Unknown | 4771 (17.2%) |
| Drinking alcohol |  |
| Daily (3 cans per day) | 2087 (7.6%) |
| Occasional | 9301 (34.1%) |
| None | 9179 (33.6%) |
| Unknown | 6738 (24.7%) |
| BMI | 23.5 [20.9, 26.5] |
| Contact history within 14 days before onset |  |
| Travel to COVID-19 countries | 432 (1.6%) |
| Contact with COVID-19 patients | 13934 (50.3%) |
| NEWS score |  |
| 0–4 | 21568 (77.4%) |
| 5–6 | 2320 (8.3%) |
| 7– | 1759 (6.3%) |
| Unknown | 2218 (8%) |
| No oxygen administration during admission | 17530 (62.9%) |
| Oxygen administration during admission | 8844 (31.7%) |
| Ventilator use during admission | 1483(5.3%) |
| Comorbidities |  |
| No comorbidity | 13659 (49.1%) |
| Any comorbidity | 14206 (50.9%) |
| Cerebrovascular Disease | 1738 (6.2%) |
| Bronchial Asthma | 1479 (5.3%) |
| COPD/Chronic Lung Disease | 1050 (3.8%) |
| Liver Dysfunction | 688 (2.5%) |
| Hypertension | 8239 (29.6%) |
| Hyperlipidemia | 4001 (14.4%) |
| Diabetes Mellitus | 4693 (16.8%) |
| Obesity diagnosed by physicians | 1686 (6.1%) |
| Moderate to Severe Renal Dysfunction/Hemodialysis | 471 (1.7%) |
| Solid Tumor/Metastatic Solid Tumor | 1062 (3.8%) |
| Leukemia/Lymphoma | 171 (0.6%) |
| Collagen Disease | 356 (1.3%) |
| HIV/AIDS | 55 (0.2%) |
| Outcome |  |
| Discharge | 20872 (74.9%) |
| Transfer to a different hospital | 3326 (11.9%) |
| Transfer to non-medical facility | 1331 (4.8%) |
| Transfer to long-term care facility | 1043 (3.7%) |
| Death | 1285 (4.6%) |

Numbers with brackets represent absolute number and percentage.
Numbers with square brackets represent median value and interquartile range.

**Table 2.**
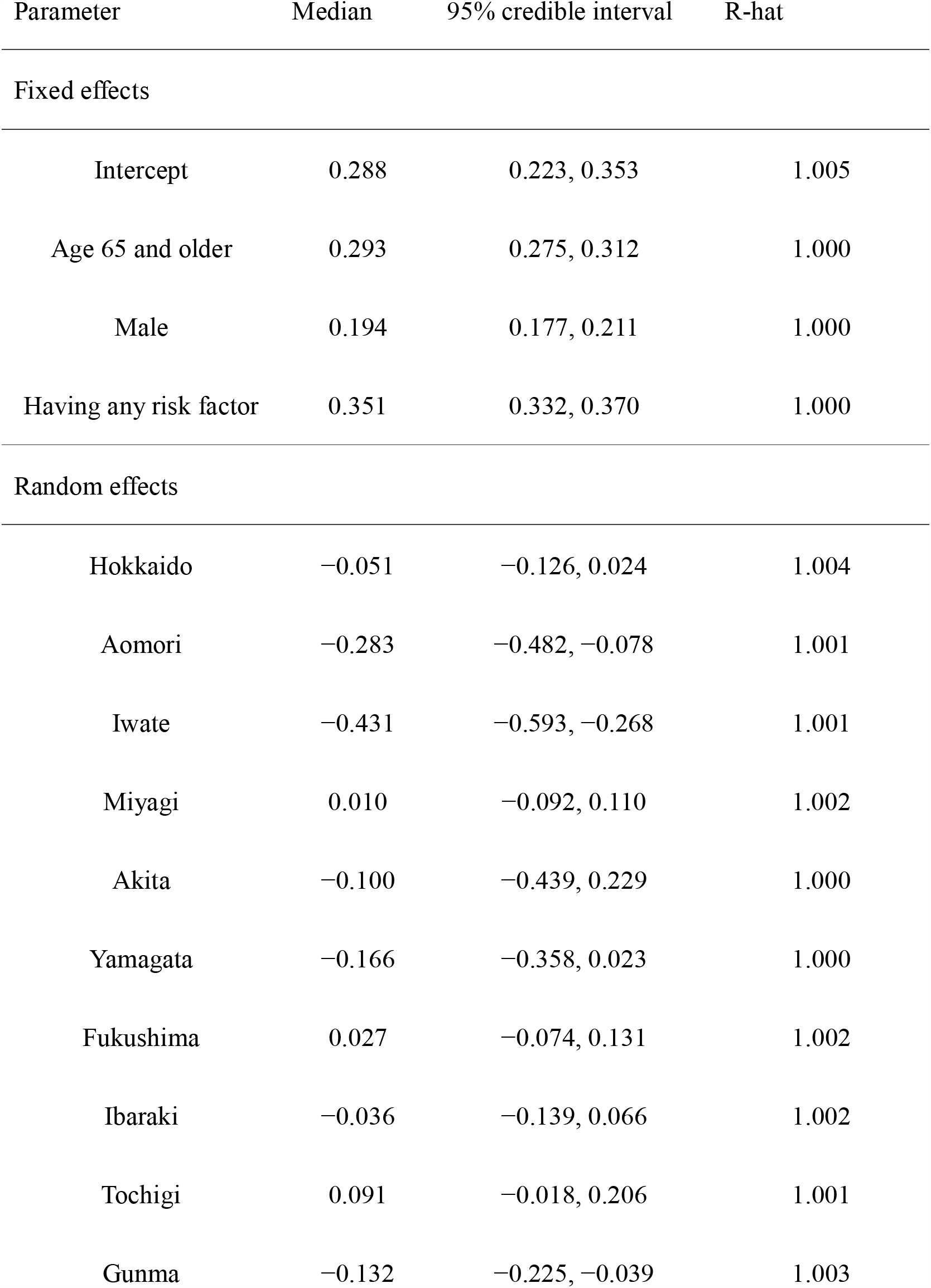
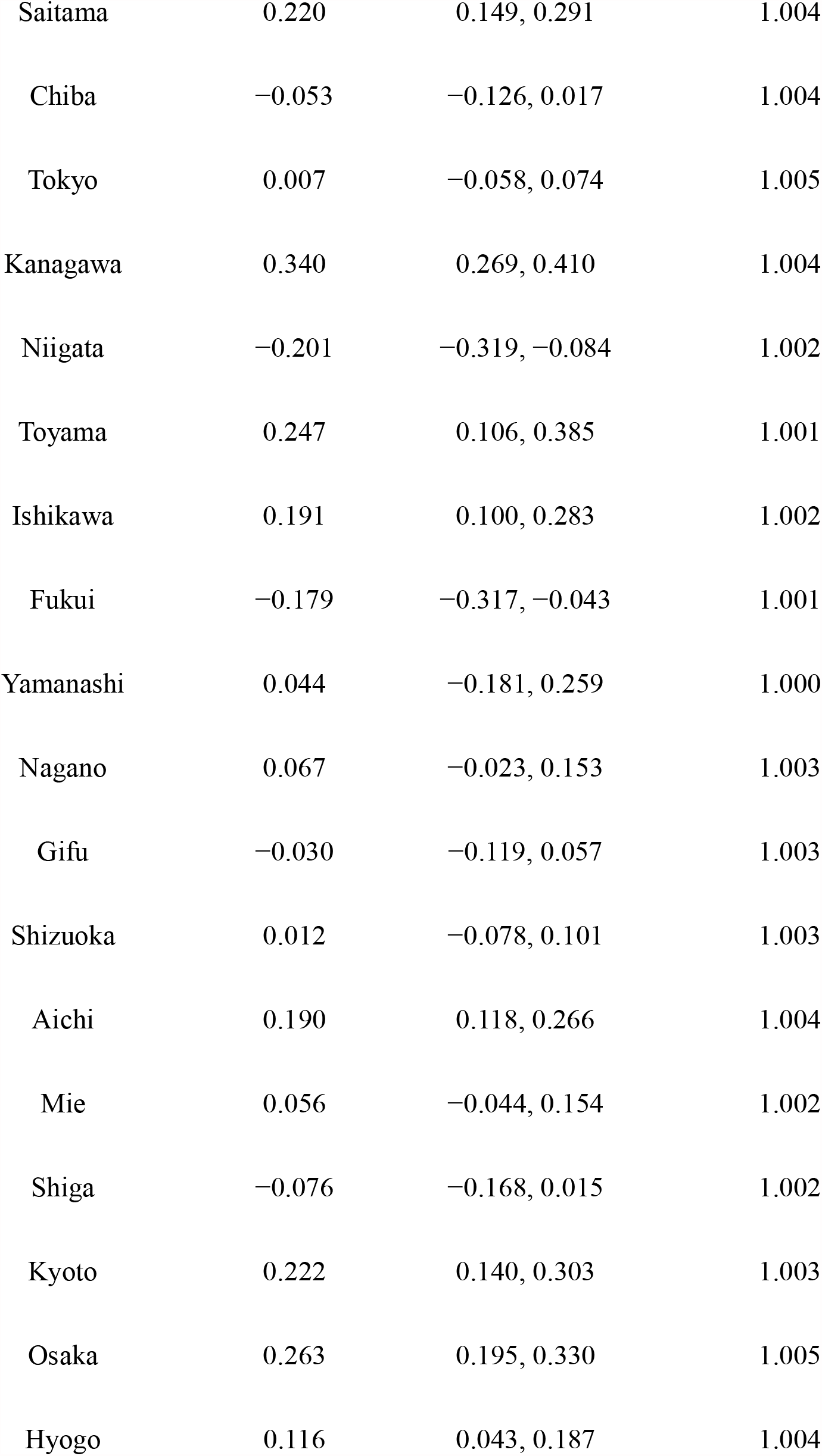
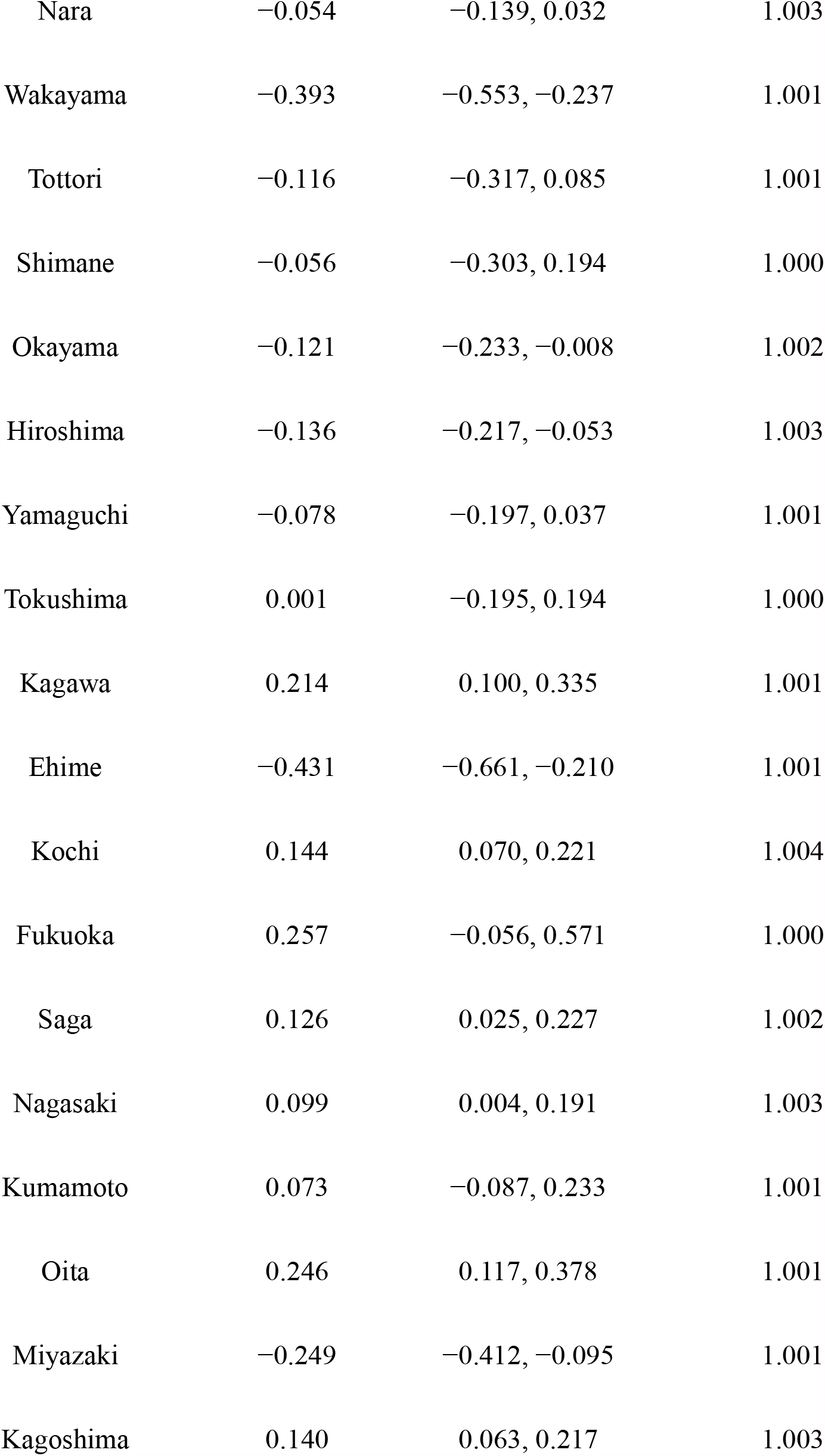
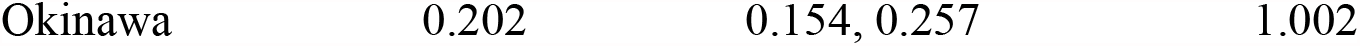
Posterior parameter distribution of hierarchical Bayesian model A.

**Table 3.** Posterior parameter distribution of hierarchical Bayesian model B.

| Parameter | Median | 95% credible interval | R-hat |
| --- | --- | --- | --- |
| Fixed effects |  |  |  |
| Intercept | 0.387 | 0.193, 0.575 | 1.005 |
| Age 65 and older | 0.293 | 0.275, 0.312 | 1.000 |
| Male | 0.194 | 0.177, 0.211 | 1.000 |
| Having any risk factor | 0.351 | 0.332, 0.470 | 1.000 |
| Number of beds<br>secured for COVID-19<br>patients | −0.004 | −0.010, 0.003 | 1.000 |
| Random effects |  |  |  |
| Hokkaido | −0.016 | −0.115, 0.081 | 1.000 |
| Aomori | −0.312 | −0.517, −0.105 | 1.000 |
| Iwate | −0.413 | −0.579, −0.249 | 1.000 |
| Miyagi | −0.030 | −0.157, 0.089 | 1.000 |
| Akita | −0.099 | −0.434, 0.230 | 1.000 |
| Yamagata | −0.182 | −0.381, 0.003 | 1.000 |
| Fukushima | 0.024 | −0.079, 0.125 | 1.000 |
| Ibaraki | -0.052 | -0.162, 0.051 | 1.000 |
| Tochigi | 0.073 | -0.042, 0.189 | 1.000 |
| Gunma | -0.153 | -0.251, -0.052 | 1.001 |
| Saitama | 0.195 | 0.112, 0.278 | 1.001 |
| Chiba | -0.070 | -0.146, 0.008 | 1.001 |
| Tokyo | 0.045 | -0.053, 0.139 | 1.000 |
| Kanagawa | 0.306 | 0.213, 0.402 | 1.001 |
| Niigata | -0.205 | -0.323, -0.090 | 1.000 |
| Toyama | 0.318 | 0.129, 0.513 | 1.000 |
| Ishikawa | 0.179 | 0.088, 0.273 | 1.001 |
| Fukui | -0.155 | -0.295, -0.006 | 1.000 |
| Yamanashi | 0.066 | -0.153, 0.295 | 1.000 |
| Nagano | 0.049 | -0.046, 0.141 | 1.000 |
| Gifu | 0.001 | -0.103, 0.107 | 1.000 |
| Shizuoka | -0.036 | -0.159, 0.089 | 1.000 |
| Aichi | 0.152 | 0.054, 0.254 | 1.001 |
| Mie | 0.041 | -0.062, 0.143 | 1.000 |
| Shiga | -0.081 | -0.174, 0.007 | 1.000 |
| Kyoto | 0.189 | 0.092, 0.291 | 1.001 |
| Osaka | 0.250 | 0.177, 0.321 | 1.001 |
| Hyogo | 0.075 | −0.028, 0.177 | 1.001 |
| Nara | −0.048 | −0.133, 0.038 | 1.001 |
| Wakayama | −0.338 | −0.529, −0.154 | 1.000 |
| Tottori | −0.033 | −0.295, 0.218 | 1.000 |
| Shimane | −0.031 | −0.286, 0.215 | 1.000 |
| Okayama | −0.137 | −0.252, −0.021 | 1.001 |
| Hiroshima | −0.169 | −0.270, −0.068 | 1.001 |
| Yamaguchi | −0.048 | −0.179, 0.081 | 1.000 |
| Tokushima | −0.011 | −0.213, 0.179 | 1.000 |
| Kagawa | 0.193 | 0.069, 0.317 | 1.000 |
| Ehime | −0.423 | −0.653, −0.201 | 1.000 |
| Kochi | 0.102 | −0.001, 0.211 | 1.001 |
| Fukuoka | 0.276 | −0.034, 0.601 | 1.000 |
| Saga | 0.147 | 0.041, 0.255 | 1.000 |
| Nagasaki | 0.109 | 0.015, 0.203 | 1.000 |
| Kumamoto | 0.092 | −0.069, 0.260 | 1.000 |
| Oita | 0.243 | 0.114, 0.374 | 1.000 |
| Miyazaki | -0.257 | -0.421, -0.101 | 1.000 |
| Kagoshima | 0.172 | 0.072, 0.265 | 1.000 |
| Okinawa | 0.200 | 0.153, 0.256 | 1.000 |

All variables other than the number of beds included as fixed effects exhibited significant associations with the NEWS score at the time of admission in both models.

Figure 1 presents the choropleth graph of the median value of the posterior distribution of prefecture-specific random intercept in model A. Figure 2 presents the choropleth graph of the median value of the posterior distribution of prefecture-specific random intercept in model B.

**Figure 1.**
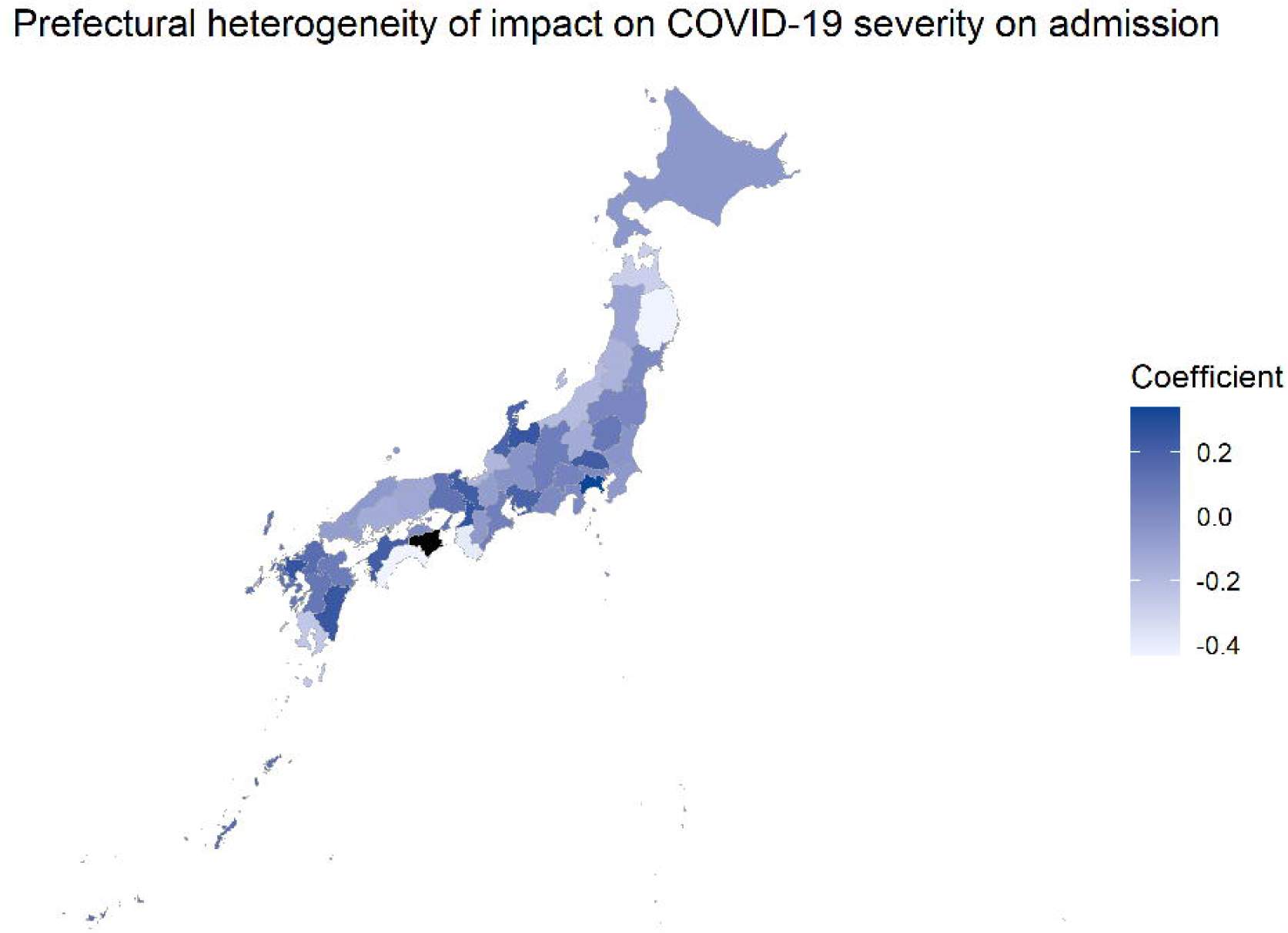
Choropleth map of prefecture-specific random effect on the severity of COVID-19 cases at the time of admission in model A. Thick colors indicate higher risk of higher NEWS score on admission. Black color (Tokushima prefecture) represents N/A as there are no registered patients in this prefecture.

**Figure 2.**
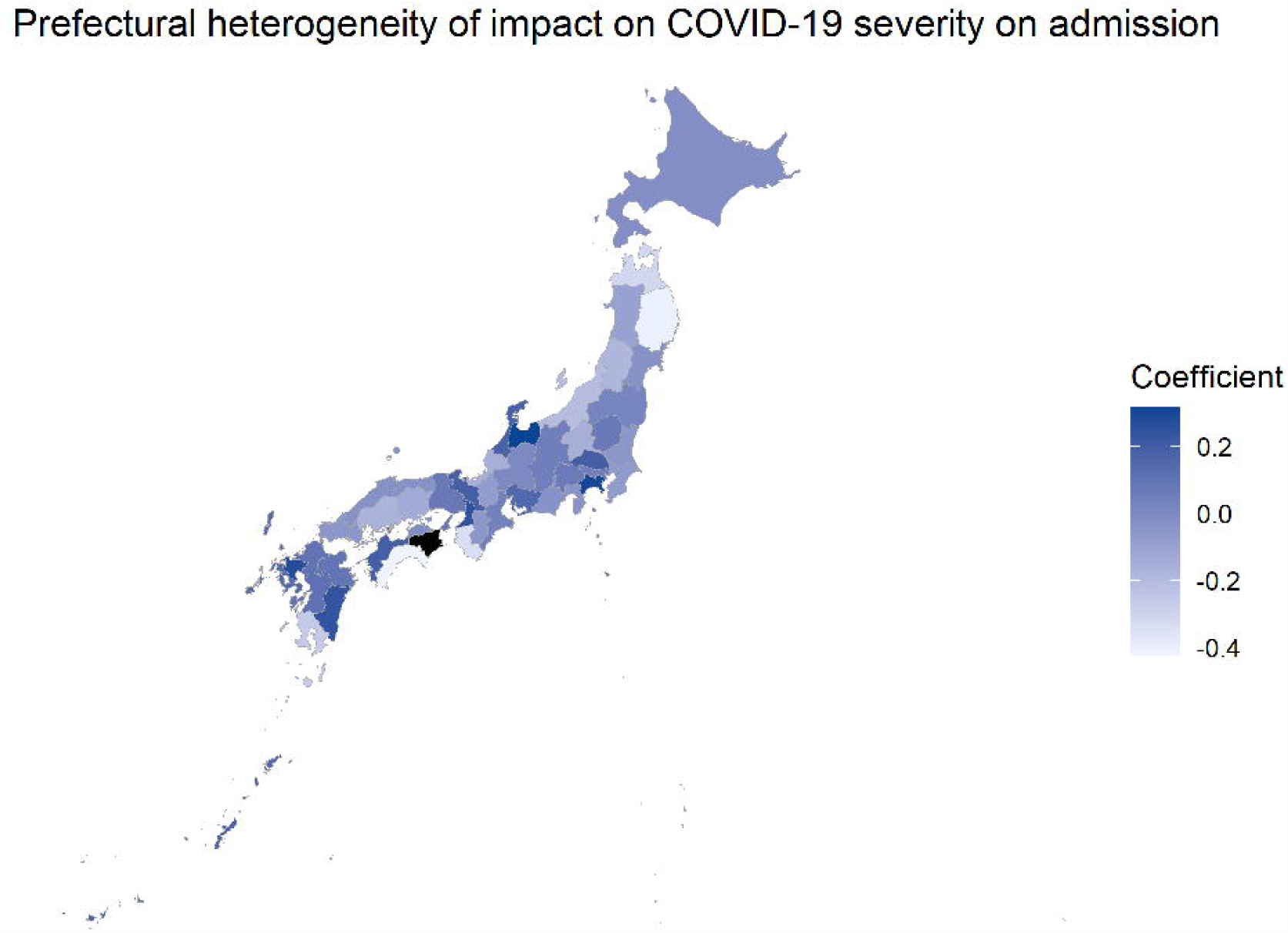
Choropleth map of prefecture-specific random effect on the severity of COVID-19 cases at the time of admission in model B. Thick colors indicate higher risk of higher NEWS score on admission. Black color (Tokushima prefecture) represents N/A as there are no registered patients in this prefecture.

Several prefectures exhibited a higher risk of higher NEWS scores upon admission, but these prefectures did not significantly change between model A and B. The Bayesian factor of model A over model B was 136988.360.

## 4. Discussion

The results revealed the existing regional heterogeneity in the severity of COVID-19 cases at the time of admission in Japan. Our results suggest that the health policy of each prefecture might have an impact on the severity of hospitalized COVID-19 cases on admission.

A wide variety was observed in the median values of the posterior distribution of random effects for the different prefectures. That of Kanagawa prefecture in model A was 0.340 [95% Credible Interval (CrI) 0.269, 0.410], and that of Iwate prefecture was -0.431 [95% CrI -0.593, -0.268], indicating a substantial discrepancy in the severity of hospitalized COVID-19 cases between these two prefectures.

The posterior distribution of random effects in model B exhibited a similar trend to that in model A. The coefficient of Kanagawa prefecture in model B was 0.306 [95% CrI 0.213, 0.402], and that of Iwate prefecture was -0.413 [95% CrI -0.579, -0.249]. Other prefectures demonstrated similar results in both models, suggesting that factors other than the number of beds secured for COVID-19 patients (e.g., prioritization policy for COVID-19 case admission and number of physicians) had an impact on the severity of COVID-19 on admission.

We should take various factors into consideration when we interpret our results. The potential factors associated with the variation of the severity on admission could be viewed from two directions, namely, the local epidemiology of COVID-19 and the COVID-19 healthcare policy in each prefecture. From the former viewpoint, a regional surge in the number of COVID-19 patients or a rapid increase in the severity of disease due to new SARS-CoV-2 variants may contribute to the change in the severity on admission.^8,14^ However, our analysis did not include patients with new SARS-CoV-2 variants as the study period was limited to before March 2021. While it is possible that COVID-19 cases due to new variants carry a greater risk of serious illness, there was a substantial regional heterogeneity in the severity of COVID-19 cases at the time of admission, as previous studies have demonstrated.^15–17^ Presumably, variants are contributing factors to the severity of COVID-19 cases, but it should be noted that other factors also have a non-negligible influence on the severity of disease.

The latter point is relevant to policymakers (and healthcare providers) in the municipalities, mainly related to the on-site indication for hospitalization that is operated in each prefecture. The policy implemented to manage the waves of infection could be affected by the total medical capacity in the area. The number of beds secured for COVID-19 patients is one of the major resources.^15,18^ The number of beds per population in each prefecture was included as one of the fixed effects in model B, and the random effect of each prefecture did not drastically change. Also, the number of physicians eligible for COVID-19 management may have an impact as this factor could create a bottleneck for patient acceptance.^19^ However, it is difficult to precisely grasp the number of physicians who are engaging in COVID-19 patient management. This is because the specialty of the medical doctor is not a good proxy for the number of physicians engaging in COVID-19 management under such an emergency period.^20^

The local government of each prefecture has established policy for the process of admission prioritization as a way of integrating the information from the regional medical situations, including the aforementioned factors, a strategy that may determine the variation in the severity on admission.^21,22^ This means that there may be differences in whether a patient can be hospitalized and how long it takes for a patient to be hospitalized, even for clinically similar cases, depending on the prefecture they belong to.

Two types of strategy for symptomatic COVID-19 patients can be assumed. The following are simplified examples: 1) moderate to severe cases are admitted to hospital, whereas mild cases are managed at home or in a dedicated facility (severity on admission will increase), and 2) all patients, whether with mild or severe disease, are admitted to hospital (severity on admission will decrease). Both strategies have their own risks and benefits, and the local government of each prefecture has adopted each strategy depending on the height of each wave of the epidemic.

The strategy adopted should be evaluated by the outcomes of COVID-19 patients, damage to the medical system for non-COVID-19 diseases, and the impact on hospital and regional healthcare economy. A previous study in China demonstrated that the number of beds and medical staff per COVID-19 patient was negatively correlated with mortality.^23^ Conversely, a recent study from Italy reported that greater access to hospitals did not affect the mortality of COVID-19 patients older than 80 years but affected that of patients younger than 80 years,^24^ suggesting that the best approach may be to change the strategy according to age group or other properties of the patients. Evaluation of the regional policy could provide important insights to construct a better healthcare system against COVID-19 and other emerging infectious diseases.

Our analysis had several limitations. First, we examined data solely from Japan; thus, it is not appropriate to generalize our results globally. Nevertheless, it is easy to imagine that, even in other countries, there would be differences in health policies among local authorities and that they might have an impact on the management of COVID-19 cases. Second, we could not include other confounding factors, such as human resources and variance of SARS-CoV-2; these aspects should be addressed in future studies.

In conclusion, our analysis revealed a possible association between prefecture and an increased/decreased risk of severe COVID-19 infection at the time of admission. In addition, the number of beds secured for COVID-19 patients in each prefecture is not a single cause of such heterogeneity; therefore, other factors could be significant for the management of COVID-19 cases in Japan. Countermeasures against COVID-19 will be more appropriate if we take these insights into consideration.

## Data Availability

The data that support the findings of this study are available upon request to the corresponding author. The data are not publicly available due to privacy or ethical restrictions.

## Acknowledgments

We thank all the participating facilities for their care of COVID-19 patients, and cooperation on data entry to the registry.

## Author contributions

Shinya Tsuzuki: Data Curation, Formal Analysis, Methodology, Visualization, Original Draft Preparation. Yusuke Asai: Data Curation, Methodology, Validation, Review and Editing. Nobuaki Matsunaga: Original Draft Preparation, Review and Editing. Haruhiko Ishioka: Original Draft Preparation, Review and Editing. Takayuki Akiyama: Data curation, Validation, Review and Editing. Norio Ohmagari: Conceptualization, Funding Acquisition, Project Administration, Review & Editing.

## Ethical approval

This study was approved by the National Center for Global Health and Medicine (NCGM) ethics review committee (NCGM-G-003494-0). Information regarding opting out of our study is available on the registry website.

## Patient consent

Informed consent was obtained in the form of opt-out on the registry website.

## Permission to reproduce material from other sources

Not applicable.

